# Endometriosis - on the intersection of modern environmental pollutants and ancient genetic regulatory variants

**DOI:** 10.1101/2025.04.15.25324328

**Authors:** Amelia Warren, Demetra Andreou, Dean Warren, Jack Wieland, Anna Mantzouratou

## Abstract

Endometriosis is a chronic gynaecological disease characterised by inflammation and can cause infertility and severe chronic pelvic pain. It is a complex disease and potentially multifactorial and tends to affect women in the same family with heritability shown to be between 47-51%. Previous studies have identified over forty single nucleotide polymorphisms associated with advanced stages of endometriosis; however, no variants have been associated with earlier endometriosis stages to date. Due to being a multifactorial condition, it is also likely that interactions with the modern environment may lead to an altered genetic susceptibility to endometriosis. The aim of this study was to identify endometriosis risk variants in selected human populations within the Genomics England database and investigate the pathways these variants could act upon within the presence of modern environmental stressors and pollutants. This study focussed on the variants of regulatory sequences of the genome likely to be impacted by environmental and genetic influencers of endometriosis. A combination of systematic analysis and data interrogation in the Genomics England 100,000 genomes project were undertaken. Participant data was extracted from participants with a diagnosis of endometriosis at various stages. Variant frequency analysis and statistical testing identified six novel regulatory variants significantly enriched in the endometriosis cohort. The strongest association was observed for two *IL-6* variants, which co-localized more frequently than expected by chance, suggesting a potential regulatory interaction. Although the sample size was limited, this study provides novel insights into how genetic regulatory variation and environmental pollutants may collectively influence endometriosis risk. The findings suggest that genetic susceptibility to endometriosis may be shaped by modern industrial exposures, with potential implications for personalised risk assessment and prevention strategies. Future research should validate these findings in larger populations and explore functional mechanisms underlying gene - environment interactions to develop targeted interventions for individuals at risk.

The study introduces a unique perspective by linking ancient Neanderthal and Denisovan genetic variants to modern disease risk in the context of environmental exposure. We propose those industrial pollutants—particularly endocrine-disrupting chemicals (EDCs)—may epigenetically disrupt immune and reproductive gene regulation, thereby amplifying disease risk in genetically susceptible individuals. This is an early-stage, hypothesis-generating study and has not yet undergone peer review. Findings should not be interpreted as clinical advice or diagnostic tools. Future research with larger cohorts and functional validation is warranted.

## Introduction

Globally, 10% of reproductive-aged women have endometriosis which is a heterogenous gynaecological disease driven by estrogen signalling (Kanellopoulos et al., 2022). Estrogen dominance can cause endometrial cells to implant outside the uterus, forming scar tissue (Angioni et al., 2020). Endometriosis can be difficult to diagnose, with Agarwal et al., (2019) finding up to eleven years can pass between symptom onset and diagnosis. Fifty percent of diagnosed women have medically reported severe pelvic pain during adolescence that went untreated (Nnoaham et al., 2011). Additional symptoms include severe menstrual cramps, heavy bleeding, with pain located in the pelvic, back, and groin, before, during, and after periods, and during intercourse (Shi et al., 2022). Infertility is also associated, as 50% of infertile women have endometriosis and 25% of endometriosis patients are infertile (Tamura et al., 2019). Limited diagnostic tools contribute to misdiagnosis and delays. While internal exams, ultrasounds, and laparoscopy can be used, current imaging techniques miss superficial lesions (Becker et al., 2022). The European society of human reproduction and embryology (ESHRE) also states diagnostics should not use biomarkers (Becker et al., 2022).

Studies have suggested endometriosis disrupts the immune system through lowered levels of check point inhibitors dampening the immune response (Santoso et al., 2020). Furthermore, Garcia-Gomez et al., (2020) and Patel et al., (2018) found estrogen dominance disrupts the immune system, triggering pro-inflammatory factors and altering immune cell function (T-cells, natural killers, macrophages). This disruption to the immune system fuels chronic inflammation, preventing cell death, and promotes the growth of endometriosis lesions (Garcia-Gomez et al., 2020). Patel et al., (2018) study found endometriosis patients show increased C chemokine production, leading to higher oxidant levels and worsened inflammation in both the lesions and surrounding peritoneal fluid.

Studies with twins by Treloar et al., (1999); Lee et al., (2013); Saha et al., (2015) show a heritability component, with these genome-wide association studies suggesting both genetic (47%) and environmental (53%) contributions to endometriosis predisposition (Saha et al., 2015).

Environmental predisposition from exposure to modern industrial pollutants and chemicals found in items like plastics, pesticides and cosmetics could potentially play a role in the development of endometriosis (Yen et al., 2019). For instance, exposure to endocrine disrupting chemicals (EDCs) imitating hormones and blocking the binding of naturally occurring hormones to their receptors may interfere with physiological processes (Hansel et al., 2024). This disruption may harm the reproductive system potentially contributing to the onset of endometriosis (La Merrill et al., 2020).

Current genome-wide studies by Sapkota et al., (2017); Zondervan et al., (2018); Rahmioglu et al., (2023) collectively identified forty-two single nucleotide polymorphisms (SNPs) linked to endometriosis. Notably, some variants were associated with pain perception and maintenance and advanced stages of endometriosis. However, none of these variants predict earlier endometriosis stages, hindering increased risk assessment accuracy and early diagnosis preventing complications such as infertility.

Despite current advancements in identifying endometriosis genes, most of the research has focused on advanced stages of endometriosis and comorbidities such as migraines and infertility, rather than the disease onset, leading to diagnosis of earlier stages and prevention of endometriosis to remain elusive. Understanding genetic risk in early stages and investigating how environmental factors interact with genes is crucial for better management and treatment of endometriosis and preventing complications such as infertility and ovarian and uterine cancer.

Genomics England (GE) has recruited over 85,000 National Health Service (NHS) patients and sequenced over 100,000 genomes from these NHS patients to enhance research in healthcare. This database has completed genomic sequences of patients with known diseases or disorders. This enables researchers to investigate genomic sequences and variants of genes and their potential impacts on known and rare diseases (The National Genomic Research Library v5.1, Genomics England https://doi.org/10.6084/m9.figshare.4530893.v7). Bournemouth University and the Department of Life and Environmental Sciences is part of the Genomics England Research Network (GERN) and has been granted access to the GE Research Environment. This enables researchers to investigate genomic sequences and variants of genes and their potential impacts on known and rare diseases.

Therefore, the aim of this study was to investigate genetic factors that may have potential interactions with environmental pollutants that can result to a combined risk to developing endometriosis. This study proposes that regulatory sequence variants in genes involved in immune modulation, inflammation, and hormone signalling contribute to endometriosis susceptibility and that these variants may be influenced by environmental pollutants, particularly endocrine-disrupting chemicals (EDCs). Specifically, we predict that non-coding genetic variants affecting the regulation of key genes may be more prevalent in individuals with endometriosis and could potentially modify disease risk through interactions with environmental pollutants.

To test this hypothesis, we analysed whole-genome sequencing data from the 100,000 Genomes Project, focusing on regulatory variants in selected genes, their frequency in an endometriosis cohort, and their potential functional consequences. This approach aims to bridge the gap between genetics and environmental risk factors, providing a more comprehensive model of endometriosis susceptibility and identifying potential biomarkers for early-stage disease detection.

## Materials and Methods

### Ethical Approval

The study received ethical approval by the BU institutional ethical approval panel, ethics ID: 45978.

### Literature Searches

To identify genomic areas and markers of interest in relation to endometriosis development and interaction with environmental factors, a comprehensive literature review was undertaken using Web of Science and PubMed. Selection criteria were applied to select relevant papers.

### Literature Selection Criteria

Inclusion Criteria: Original studies focusing on genomic/genetic analysis, or genome wide association design., Only human participants (no other species)., Patients with a diagnosis of endometriosis for at least a year., Patients aged between 18 and 43 years at the time of recruitment.

Exclusion Criteria: Review studies., Studies including participants with other types of female infertility., Participants without an endometriosis diagnosis., Participants with additional illnesses and disease which could affect the outcome of the results.

A literature search was conducted to investigate environmental factors that are adversely associated with endometriosis development risk. A review of how these factors affect signalling pathways and the genes involved in them was conducted. The key words used for environmental factors were; “endometriosis and “exposure to endocrine disrupting chemicals”, “endocrine disrupting chemicals”, “exposure to pesticides”, “pesticides”, “personal care products”, “cosmetics”, “exposure to heavy metals”, “heavy metals”, “exposure to radiation”, “radiation”, “exposure to toxins”, “toxins”, “chemicals”, “plastics”, “exposure to pollution”, “pollution”, “exposure to air pollution”, “air pollution”, “exposure to water pollution”, “water pollution”. This search yielded sixty-four papers and 236 papers were excluded. A PRISMA flow chart presented in figure one shows the screening prosses of the primary literature search.

**Figure 1:**
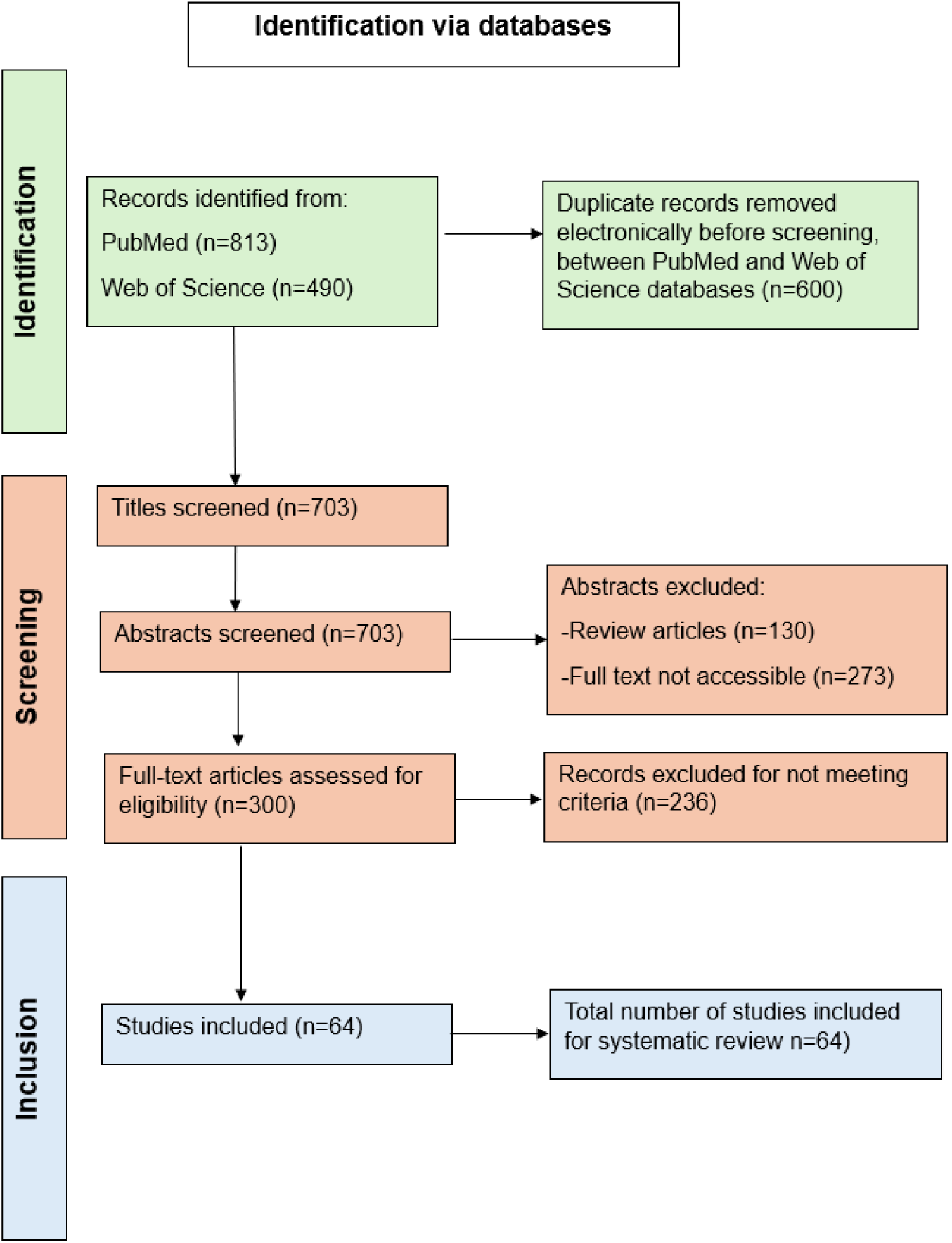
The PRISMA flow chart used to select literature for the impact of the environment on endometriosis risk.

This literature review identified EDCs as the most significant environmental factor to endometriosis. Therefore, this study focused on EDC’s exploring their properties, structure, uses, mechanisms, signalling pathways effected by exposure of EDCs and their association with genetic susceptibility to endometriosis.

A secondary literature search was conducted to identify genomic areas of interest and genetic markers involved in endometriosis development. The key words used in this search were “endometriosis and “polymorphism” or “SNP” or “genetic polymorphism” or “variants” or “locus” and “GWA” or “Genome-wide” or “Genome wide” or “Genetic association study”. This yielded 166 papers to be included and 2476 papers to be excluded. A PRISMA flow chart presented in figure two shows the screening prosses of the secondary literature search.

**Figure 2:**
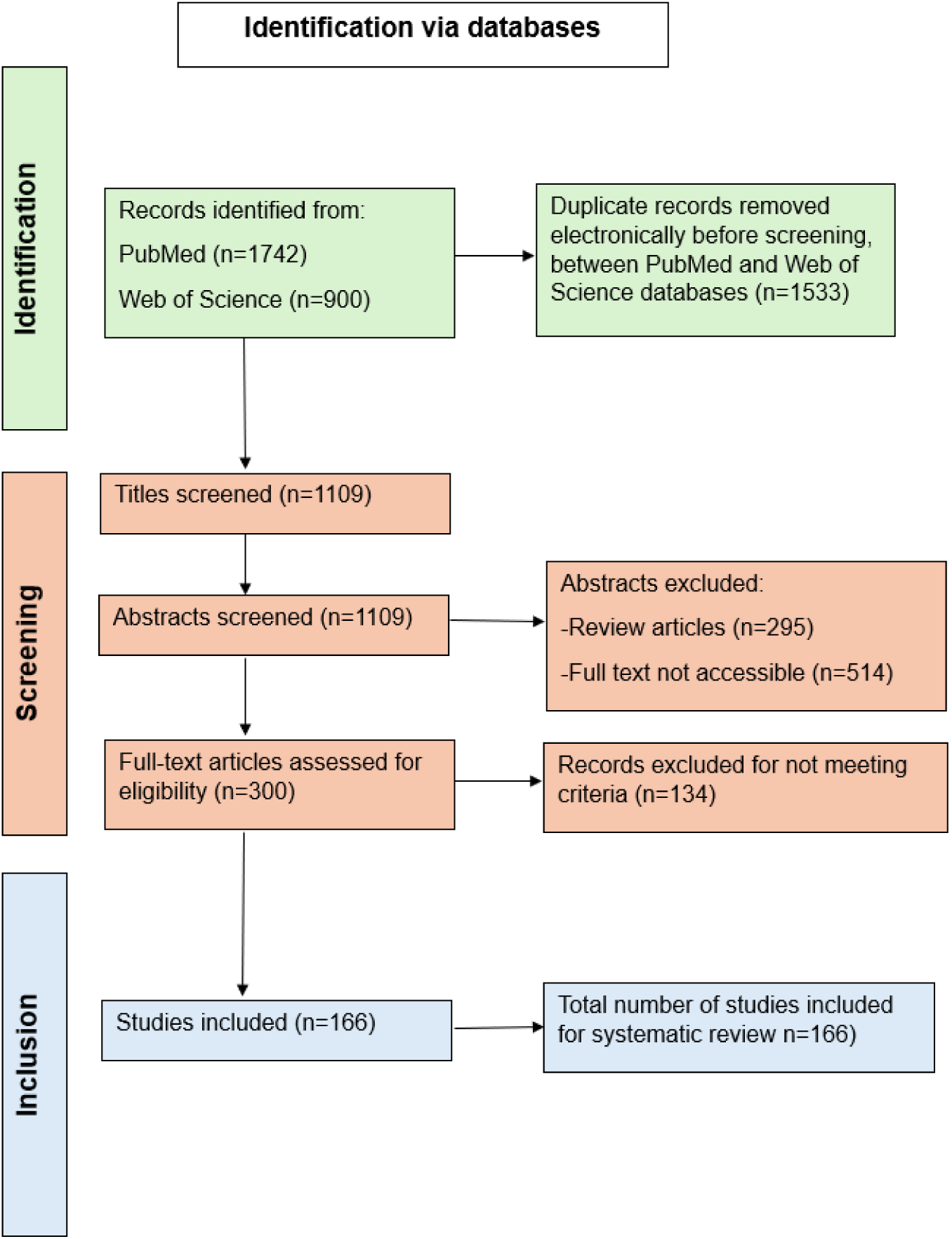
PRISMA flow chart created for the screening process to select literature on genomic areas of interest in relation to endometriosis development.

This literature search identified fifty-seven genes of interest in relation to endometriosis risk susceptibility, and from this, five genes were chosen to investigate further due to published studies demonstrating these were affected by the action of EDCs. These were chosen through identifying the main function of the gene and where it is expressed using the national library of medicine (NIH) (https://www.nlm.nih.gov/). The pathways of the genes and their associated variants were identified using Reactome (Gillespie et al., 2022). The criteria for this selection are outlined below:

- Literature has shown EDCs are associated with disruption to the gene; this could be through altered expression or epigenetic alterations such as cytosine demethylation and modification of histones (Alavian-Ghavanini, and Rüegg, 2018).
- Literature has shown EDCs effects are within the signalling pathways that selected genes are involved in. Leading to an alteration to the pathway and disrupting the homeostasis (Lauretta et al., 2019).
- The pathways associated with the gene are seen to be associated or involved in endometriosis onset or progression.
- The selected genes are expressed within tissues associated with endometriosis implant locations.
- The main function of the gene when disrupted potentially leads to development of endometriosis.

### 100,000 Genomics England Database

The GE 100,000 genome database was used in this study as a part of an ongoing larger project “Genomic and chromosomal instability sequence markers in relation to fertility, early pregnancy, and cancers of the reproductive tissues” Project ID 645 which included domains of Ovarian Cancer GERN and Endocrine and Metabolism GERN to obtain participants and analyse genomic data.

### Study population from the Genomics England Database

Participants were chosen using the GE Rare Disease Programs GRCh38 Participant Explorer by searching for clinical diagnosis of endometriosis.

### Participant Selection Criteria

Inclusion Criteria: Female participants aged 18–43 years at the time of recruitment., Diagnosis of endometriosis recorded in medical history., Availability of whole-genome sequencing (WGS) data., Inclusion of participants with endometriosis-related infertility and/or ovarian chocolate cysts.

Exclusion Criteria: Individuals not assigned female sex at birth., Participants over the age of 43 (to minimize confounding by menopause-related genetic changes)., Presence of additional ovarian pathology, chromosomal abnormalities, haematological disorders, or other reproductive tract malignancies., Diagnosis of diabetes, immunological disorders, or hormonal conditions that could confound genetic associations., Body mass index (BMI) outside the range of 18.5–30 kg/m² to minimize the impact of metabolic confounders. The final study cohort included 19 individuals meeting these criteria.

The stage of endometriosis was not stated in the Participant Explored database, so endometriosis stages were estimated according to the American Society of Reproductive medicine (Zhang et al., 2019), using additional clinical information available in the patient’s profile. This included the locations of endometrial implants and/or lesions and procedures or investigations carried out in relation to endometriosis with table one depicting the study criteria to estimate the stage of endometriosis patients in this study cohort. This led to four patient cohorts for stages one to four.

**Table 1:** Criteria used in this study to estimate the patient’s endometriosis stage according to the American Sociated of Reproductive medicine (Zhang et al., 2019).

| Stage | Clinical information to diagnose endometriosis stage |
| --- | --- |
| Stage 1 | Minimal surgeries such as examination of uterus and laparoscopic approach to abdomen, with one or two locations of endometriosis within the ovary, pelvic peritoneum, uterus or rectovaginal septum and vagina. |
| Stage 2 | Had two or more locations of endometriosis within the ovary, pelvic peritoneum, uterus or rectovaginal septum and vagina and minimal surgeries such as examination of uterus and laparoscopic approach to abdomen, including removal of lesions. |
| Stage 3 | Multiple locations of endometriosis, chocolate cysts and multiple surgeries including cauterisation of organs, |
|  | endoscopic freeing of adhesions and extirpation of ovaries. |
| Stage 4 | All locations of endometriosis including possible endometriosis located in the intestine in some cases, chocolate cysts and procedures of endoscopic resection, endoscopic destructions, repair of obstetric lacerations, drainage of ovarian cysts and endoscopic extirpation. |

### Determining genes and variants related to endometriosis risk

The GE Interactive Variant Analysis (IVA) workspace was used to identify potential genes and variants linked to the risk of developing endometriosis. This workspace allows searches for variants found in the selected genes in the participants in this study, through selecting for genes, consequences and frequency in patient IDs and family genotypes. Each of the five genes selected to investigate further were searched in each participant and single nucleotide variations (SNVs), and insertion-deletion mutations (INDEL’s) were collected.

The regulatory sequences introns, upstream and downstream sequences were investigated in this study rather than the coding sequences. Investigating the regulatory sequence variants and their impact on gene expression was chosen due to environmental pollutants having a higher possibility of acting upon the expression of the targeted genes rather than the protein itself (You et al 2021). Also, modern industrial pollutants may also influence the variants which may otherwise not be disadvantageous. This highly informed and targeted approach lead an efficient and more detailed investigation of flagged genomic sequences.

### Statistical analysis

Heatmaps were created using GraphPad Prism version 10 to visualise variants found in multiple stages and variants. These were then compared to the whole population within GE and the endometriosis cohort, which was subsequently followed with a χ^2^ goodness of fit test for individual variants with Fisher’s combined probably and small sample corrections and checks relevant to data discovery which allowed some flexibility. To account for multiple hypothesis testing, a Benjamini-Hochberg (BH) false discovery rate (FDR) correction was applied to all p-values, controlling for false positives while maintaining statistical power.

To confirm that the enrichment of significant variants was specific to the endometriosis cohort, a randomly selected group of 19 individuals from the Genomics England database (with no endometriosis diagnosis) was analysed using the same variant screening process. A Fisher’s combined probability test was conducted to compare the endometriosis cohort, random control sample, and Genomics England total population, providing an overall measure of statistical significance across variant frequencies. Variants identified as statistically significant were assessed for co-localization effects to determine whether they exhibited non-random clustering within the endometriosis cohort. Linkage disequilibrium analysis was conducted to assess the correlation between regulatory variants associated with endometriosis. Pairwise LD values (D’ and r²) were calculated for rs34880821 and rs2069840 in *IL-6* and rs806372 in *CNR1*, using data from the 1000 Genomes Project across multiple populations. LDpair and LDpop from LDlink was used to determine linkage strength, with comparisons across African, East Asian, European, South Asian, and Admixed American populations. The results were analysed to evaluate population-specific evolutionary pressures and potential functional implications for immune regulation and pain sensitivity.

### Regulatory Sequence analysis

In order to find the impact of statistically significant variants found in this study an extensive search of the variants rsID was undertaken using ClinVar, dbSNP, ensemble, UCSC, and String, the workflow for this search is shown in figure three.

**Figure 3:**
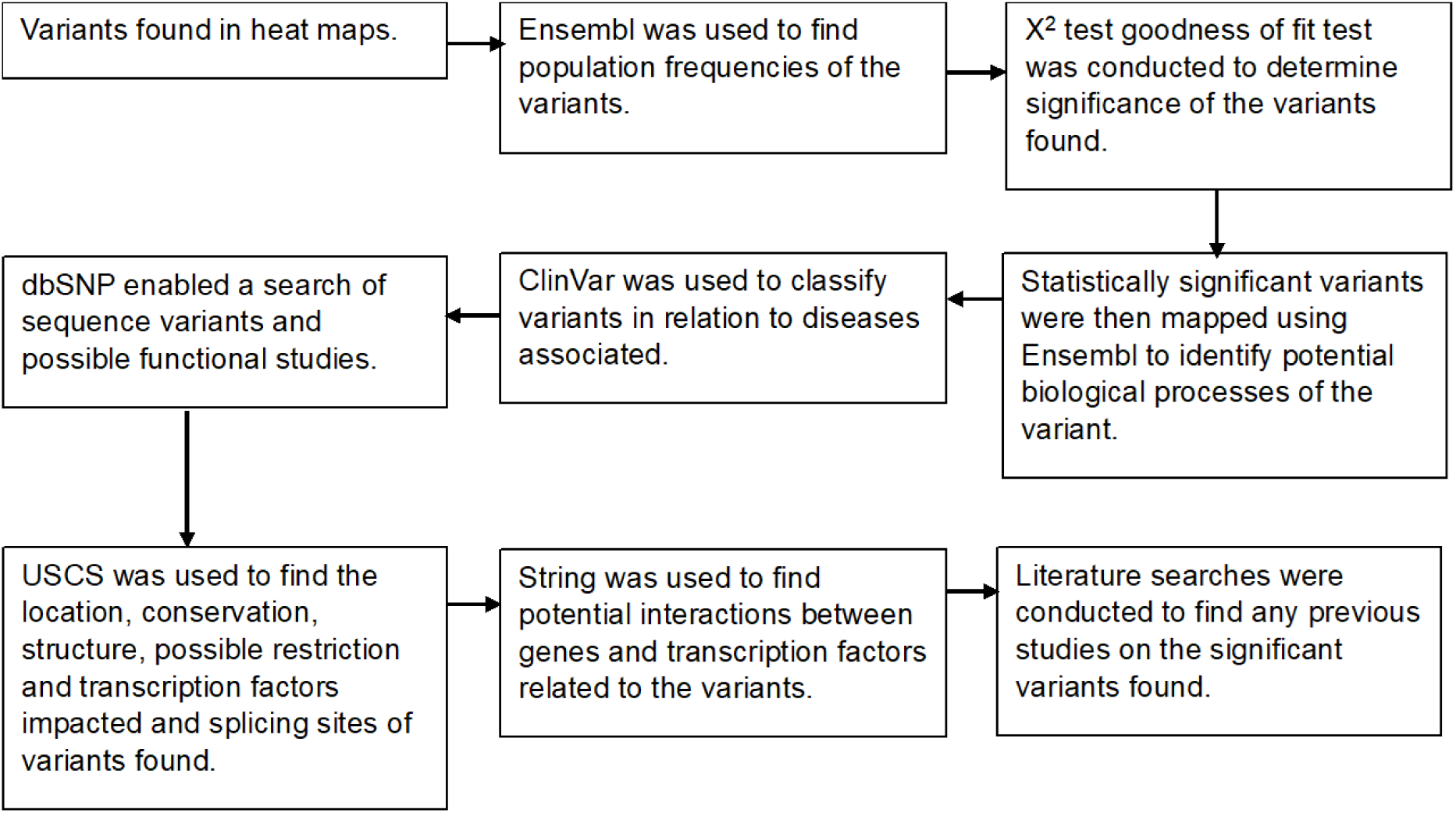
Workflow of methods followed for finding the impact and regulatory sequences of statistically significant variants.

An extensive analysis was conducted using UCSC and previous literature on other hominoids and conservation of the statistically significant variants, this can help to identify differences which can lead to disease predisposition (Jayapalan et al., 2016). All results are reported with adjusted p-values, and statistically significant associations are discussed in the context of biological plausibility and environmental interactions.

## Results

### Genomic findings of five selected genes in study participants

Through using the literature searches and consulting the criteria, the five genes chosen to be investigated further were *IDO1(*Indoleamine 2,3-dioxygenase 1), *IL-6 (*Interleukin 6), *CNR1 (*cannabinoid receptor 1), *TACR3* (tachykinin receptor 3) and *KISS1R (*KISS1 receptor).

### Significant variant findings

Ten variants were found to be potential contributors of endometriosis when comparing the study population and GE total population. When comparing the variants found in Ensembl and GE total populations, 40% of the variants had 0% frequency in Ensembl general population but were present in GE general population. This is potentially due to GE participants having a higher percentage of genetic mutations and variants through recruiting participants with known rare genetic conditions or specific diseases.

After conducting a χ^2^ goodness of fit test for each of the ten variants found using the study cohort population frequency and GE total population frequency, variants rs806372, rs76129761, rs2069840, rs34880821, rs933717388, and rs72643906 were found to be statistically significant.

A BH test was conducted from the χ^2^ goodness of fit test. This test shows the variants come up as significant in the endometriosis cohort. When observing co-occurrence with other significant variants rs2069840 and rs34880821 showed the strongest co-occurrence shown in eight individuals in the endometriosis cohort.

Whereas rs806372 and rs933717388 shows moderate co-occurrence in three individuals. However, rs76129761 and rs72643906 rarely occurs with other variants. When conducting a co-localisation analysis, rs2069840 and rs34880821 shows a strong co-localisation effect. The expected frequency for co-localisation for the variants is two participants in the cohort of nineteen (10.5%), based on P(co-localise) = 0.316×0.263=0.083. In the endometriosis group eight individuals had both variants (47.4%), which is significantly higher than expected at p=0.0001, showing a strong co-localisation effect. However, in the random group this was seen in six individuals (31.6%), this was also higher than expected at p=0.0110 but is less pronounced than the endometriosis group. This shows the co-localisation of rs2069840 and rs34880821 occurs much more frequently than would be expected by chance in both groups. Furthermore, both variants individually and together show significant enrichment in the endometriosis group. This suggests a potential biological interaction between these variants that may be particularly relevant to endometriosis. Table two shows the variant profiles for the significant variants found within the endometriosis cohort.

**Table 2:** Variant profiles for the significant variants found within this cohort (created using UCSC (2023)).

| Variant | Associated gene. | Variant type/consequence. | Chromosome location. | Potential transcription factor (TF) binding site affected (if applicable). | Number of publications. | Other significant features. | Pollutant interference. | P-value from $\chi^2$ goodness of fit | BH-corrected p-value |
| --- | --- | --- | --- | --- | --- | --- | --- | --- | --- |
| rs72643906 | *IDO1* | A > G<br>Downstream variant | Chr8:39779157-399779157 | *ZIC2* (Zic family member 2) | None | Highly conserved in mammals and vertebrates | BPA and TBBPA (Tetrabromobisphenol A) can decrease methylation of *IDO1* 's mRNA<br>and DNA<br>(Merrill et al., 2023), and TCDD can increase IDO1 expression levels (Matta et al., 2021).<br>Exposure to BPA has been shown to decrease *ZIC2* expression (Brown, 2009). | 0.00175665990877229 | 0.006 |
| rs93371<br>7388 | *KISS1R* | C > G<br>Intron<br>variant | Chr19:91<br>2365-912365 | *ZNF707* (Zinc<br>finger protein 707)<br>and *ZBTB11* (Zinc<br>finger and BTB<br>domain-containing<br>protein 11) | None | Conserved<br>in humans | DEHP<br>exposure<br>influences<br>regulators of<br>HPGA by<br>regulating<br>GnRH<br>(Gonadotropin<br>hormone-<br>releasing<br>hormone)<br>release<br>altering<br>*KISS1R* expression<br>(Carli et al.,<br>2022). | 0.0175309507<br>08949900 | 0.036 |
| rs34880<br>821 | *IL-6* | G > A<br>Intergenic | Chr7:227<br>75450-2277545<br>0 | None | 3 | Conserved<br>in humans<br>Methylation<br>site.<br>Neandertal<br>variant. | Exposure to<br>TCDD can<br>increase the<br>expression of<br>*IL-6* (Matta et<br>al. 2021). | 0.0060425862<br>45540780 | 0.024 |
| rs20698<br>40 | *IL-6* | C > G<br>Intronic | Chr7:227<br>68572-2276857<br>2 | None | 12 | Conserved<br>in rhesus,<br>mouse,<br>and dogs. | Exposure to<br>TCDD can<br>increase the<br>expression of | 0.0200596067<br>54631400 | 0.040 |
|  |  |  |  |  |  |  | *IL-6* (Matta et al. 2021). |  |  |
| rs76129761 | *CNR1* | CTCT ><br>CT<br>Intron | Chr6:88860156-88860156 | *ZNF701* (Zinc finger protein 701), *SOX4* (SRY-box transcription factor 4), *SOX6* (SRY-box transcription factor 6), *FOXD3* (Forkhead box D3) and *SOX11* (SRY-box transcription factor 11). | None | Conserved in human, rhesus, mouse, and dogs. | Exposure to DiNP and BPA significantly increased the expression of *CNR1* (Forner-Piquer et al., 2018).<br>Exposure to BPA can upregulate *SOX4* expression | 0.023331593081759400 | 0.046 |
|  |  |  |  |  |  |  | (Kang et al., 2014), SOX6 and SOX11 (Lichtensteiger et al., 2021).<br>Exposure to BPA has been shown to decrease *FOXD3* levels (Baba et al., 2009). |  |  |
| rs80637<br>2 | *CNR1* | C > G<br>Intron | Chr6:888<br>46563- | *SOX12 (SRY-box*<br>*transcription factor*<br>*12).* | 3 | Conserved<br>in humans | Exposure to DiNP and BPA significantly | 0.0033128175<br>23356740 | 0.018 |
|  |  |  | 8886656<br>3 |  |  | and<br>rhesus.<br>Splicing<br>site; SIB<br>locus ID:<br>NC-000006-1470. 3<br>alternative<br>transcripts<br>within<br>intron 1<br>(6q15) of<br>*CNR1*. | increased the<br>expression of<br>*CNR1* (Forner-Piquer et al.,<br>2018).<br>DEHP can<br>increase<br>*SOX12*<br>expression<br>levels (Chou et<br>al., 2019). |  |  |
|  |  |  |  |  |  | Denisovan<br>variant |  |  |  |

When comparing the six significant variants in the endometriosis cohort compared to the random sample cohort and the total GE population, each of the variants are shown to be significantly higher in the endometriosis group. Except rs76129761 which is shown to be higher in the random sample cohort.

From the fisher’s combined probability test between the endometriosis cohort and the random sample compared to the GE total population a combined χ^2^ statistic; 44.37, with a p-value of 0.000013 (p < 0.001) showed strong evidence in overall frequency difference between the endometriosis cohort and the GE total population across all endometriosis variants. This suggests that the genetic profile of the endometriosis cohort significantly deviates from the general population, potentially indicating a unique genetic signature associated with endometriosis. Furthermore, the combined χ^2^ statistic for the random sample cohort; 12.23, with a p-value of 0.427, showed no significant difference between the random sample cohort and the GE total population across all endometriosis variants. This lack of significant deviation suggests that the Random Sample’s variant frequencies are representative of the GE Total Population, serving as a control group.

A summary of what has been found in this results section has been provided in figure four.

**Figure 4:**
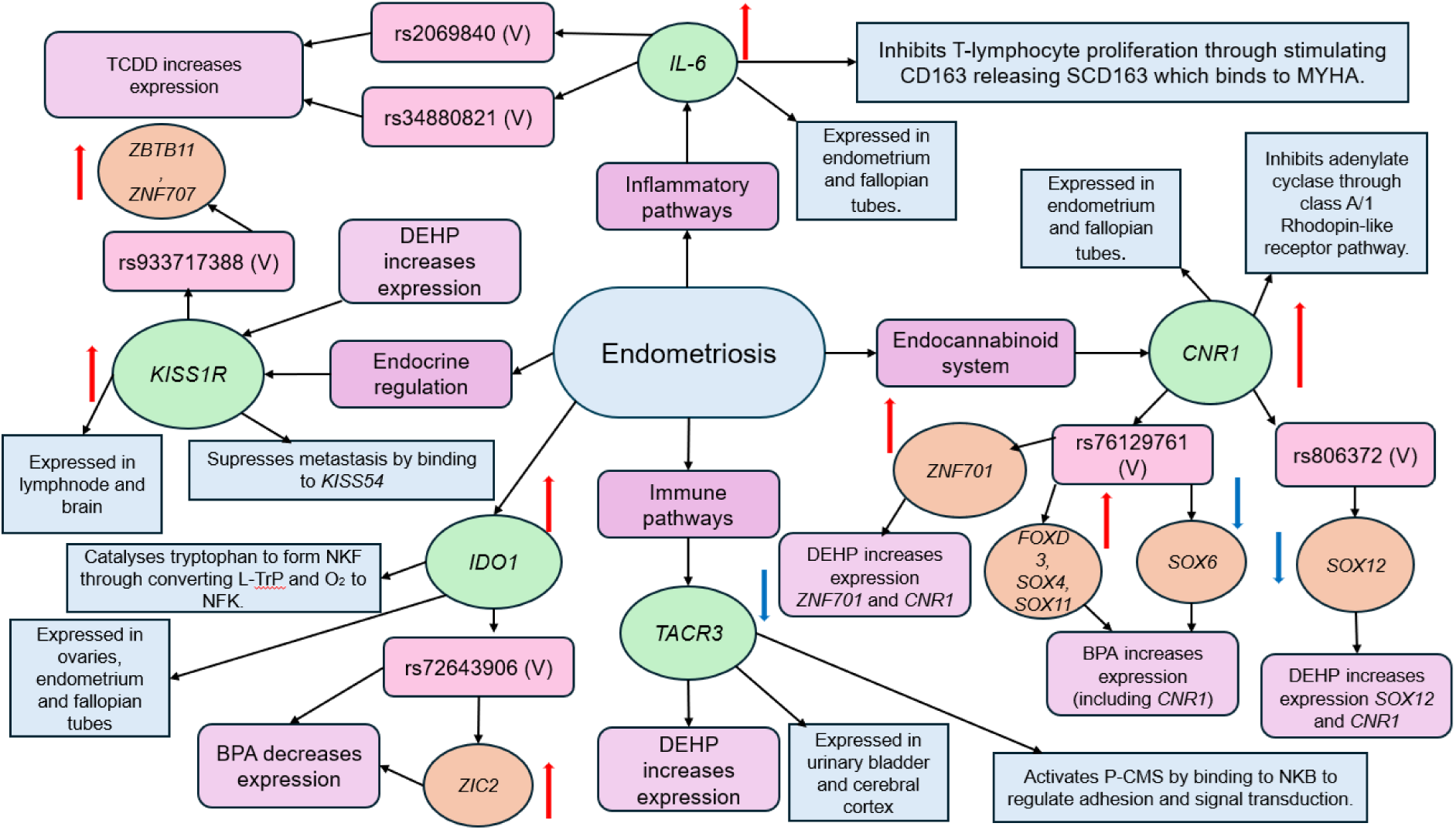
A visual representation of the hypothetical relationships between the identified variants and environmental pollutants, and how they could relate to endometriosis onset and progression. The red arrows represent potential increased expression levels of genes, with blue arrows represent potential decreased expression of genes and transcription factors in endometriosis patients if selected the variants were to be present and disrupt expression. Green circles represent genes, orange circles represent transcription factors, blue squares represent properties of each gene, light pink boxes represent pathways, light purple boxes represent endocrine disrupting chemicals effect on genes and transcription factors and light red boxes represent variants.

### LD analysis

LD analysis revealed strong linkage between Neanderthal-derived *IL-6* variants (rs34880821 and rs2069840) (depicted in figure 5), in East Asians (D’ = 1.0, r² = 0.9662), suggesting selective retention and potential immune regulation effects. Europeans showed moderate LD (D’ = 0.9234, r² = 0.5794), while Africans had weak linkage (D’ = 0.8752, r² = 0.4823), likely due to the absence of Neanderthal introgression.

**Figure 5:**
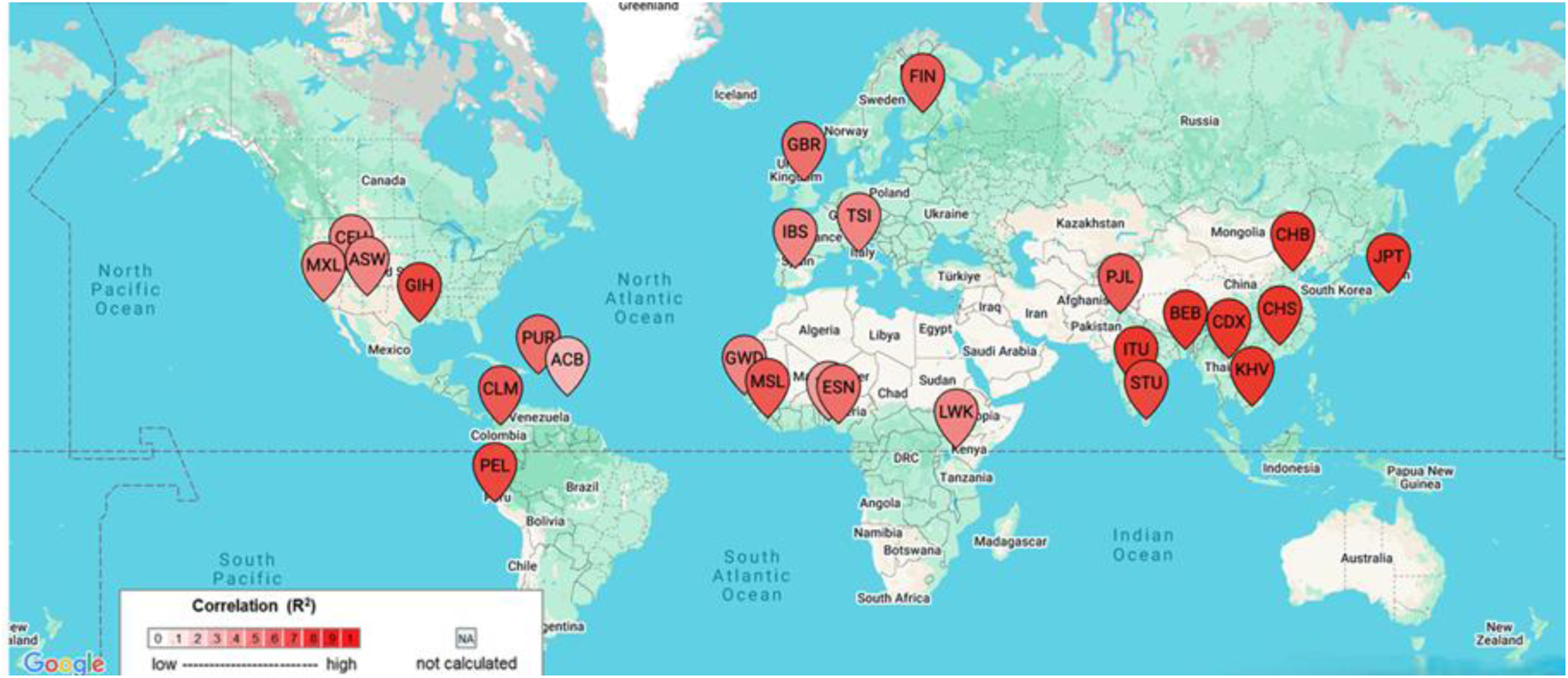
LD analysis of variants rs34880821 and rs2069840.

The Denisovan-influenced *CNR1* variant (rs806372) exhibited moderate LD in East Asians (D’ = 0.8004) but weak LD in Europeans (D’ = 0.4607) and South Asians (D’ = 0.1459), suggesting population-specific evolutionary pressures. African populations had negligible LD for both variant pairs, supporting the hypothesis that these associations arose post-migration due to archaic human introgression.

This study identified six novel regulatory variants enriched in the endometriosis cohort, with *IL-6* variants showing strong co-localization, suggesting a role in inflammation. Variants in *CNR1*, *IDO1*, and *KISS1R* were also significantly associated, with functional analysis indicating potential disruptions in transcription factor binding and immune regulation. Several variants overlapped with regions affected by endocrine-disrupting chemicals, highlighting potential gene-environment interactions in disease susceptibility. While limited by sample size, these findings provide new insights into how regulatory genetic variation and modern environmental exposures may collectively influence endometriosis risk, warranting further validation in larger studies.

## Discussion

Many sufferers of endometriosis are subject to misdiagnosis and long diagnosing times, through a lack of understanding of risk factors in earlier endometriosis stages. To obtain a better understanding of genetic factors predisposing endometriosis development, two literature searches and interrogation of the GE 100,000 genome database were carried out.

Using a highly targeted multilevel approach five genes have been characterised as potential targets in the developing endometriosis pathway, containing variants or having altered expression levels, with five variants found as highly significant and one as moderately significant in our cohort population when compared to the GE total population. This builds on the work conducted by Sapkota et al., (2017), Zondervan et al., (2018) and Rahmioglu et al., (2023) who collectively found forty-two SNPs linked to endometriosis.

### IDO1

The downstream variant rs72643906 of *IDO1* was significantly higher in the study cohort, which may potentially influence endometriosis development through altering expression levels of *IDO1*. UCSC, (2023) data found rs72643906 to alter the motif of the transcription factor Zic family member 2 (*ZIC2)*, which regulates tissue expression (Luo et al., 2015). Although, rs72643906 is found to be very rare, UCSC, (2023) data has shown this variant to be highly conserved in mammals and to be associated with an elevated risk of COVID-19. Because of this, it can be hypothesised that rs72643906 is disadvantageous to the immune system. This can be validated through Brooks et al., (2016) study finding *IDO1* expression to be increased by immune system activation. It is possible to hypothesise rs72643906 modifies *ZIC2* motif preventing the transcription factor from binding increases *IDO1* and *ZIC2* expression levels. However, this effect is potentially reversed through exposure to BPA decreasing expression levels of *IDO1* and *ZIC2* when it is introduced through the environment into the pathway (figure four). Increased *IDO1* expression levels have been associated with endometriosis risk in research conducted by Mei et al., (2013) and Kong et al., (2021) finding elevated expression of *IDO1* in endometriosis patients.

### CNR1

This research showed two significant variants of *CNR1,* rs76129761 a deleterious variant which has not been referenced in any previous publications and rs806372, referenced in previous literature, however, the literature did not have relevance to endometriosis. These variants were found to be frequent within the study population, and rs76129761 was found to be highly conserved within mammals and rs806372 had a potential splicing site implicating three alternative transcripts within intron 1 (6q15) of *CNR1* (UCSC, 2023). Due to being frequent in the study population and having a splicing site, it is possible the variants affect *CNR1* expression levels potentially leading to endometriosis development. Research by Allam et al., (2022), and Bouaziz et al., (2017) found increased levels of *CNR1* in endometriosis patients when compared to controls.

UCSC (2023) data showed rs806372 to be a Denisovan variant, which is prevalent in east Asian populations, who have a high prevalence of endometriosis (15.4%), compared to European (1.4%) and general (4.44%) populations (Parazzini et al., 2020). This shows that the variant might be well established in populations where Denisovans and sapiens interacting being randomly selected until the onset of EDCs in our modern society (Zhang et al., 2021). This impact may have been increased by DEHP, DiNP and BPA which have previously been shown in this paper to increase *CNR1* expression and potentially in turn endometriosis risk (Ernst et al., 2020; Forner-Piquer et al., 2018).

The *CNR1* variant rs806372 may potentially modify the *SRY-box transcription factor* (*SOX) 12* motif (UCSC, 2023) preventing the transcription factor from binding and causing increased expression of *CNR1* and decreased *SOX12* levels. If DEHP is introduced to the pathway through exposure from the environment the expression of *SOX12* and *CNR1* is increased an observation supported by the expression studies (Chou et al., 2019; Ernst et al., 2020).

The significant variant rs76129761 of the *CNR1* gene potentially changes the motif of five potential transcription factors. All the transcription factors Zinc finger protein 701 (*ZNF701)*, *SOX4*, *S0X6*, Forkhead box D3 (*FOXD3)* and *SOX11* when in the presence of rs76129761 potentially increase expression levels alongside increased *CNR1* expression. Furthermore, when exposed to BPA this consequence is potentially further increased in the pathways including *SOX4*, *SOX6*, and *SOX11* (Lichtensteiger et al., 2021; Kang et al., 2014). However, in the presence of BPA *FOXD3* was decreased (Baba et al., 2009), although there was no literature available for *ZNF701* and other EDCs.

### KISS1R

The intronic variant rs933717388 of *KISS1R* was found to be significantly higher in the study population and potentially novel variant due to no literature or records referencing this variant. Data obtained from UCSC (2023) found rs933717388 to lie on the binding sites of Zinc finger protein 707 (*ZNF707)* and Zinc finger and BTB domain-containing protein 11 (*ZTB11)* and modifies the motifs, potentially preventing the transcription factors from binding. This can alter expression of KISS1R, specifically *ZTB11* is a silencing transcription factor, which means potentially increased *ZTB11* levels as the transcription factor might not be binding as readily resulting in increased *KISS1R* expression levels. Therefore, it may be hypothesised that rs933717388 causes an increased expression of *KISS1R* which may lead to dysregulation of the HPGA and reproductive regulation, while increasing metastasis of endometriotic cells outside of the uterus (Xie et al., 2022; Gillespie et al., 2022). This is supported by Blasco et al., (2020) finding abnormal and heightened expression levels of *KISS1R* in endometriosis patient granulosa cells when compared to controls. Within both pathways, *KISS1R* and transcription factors expression levels are increased, with exposure to DEHP increasing *KISS1R* expression levels further, although there is not literature for DEHP effect on *ZNF707*, or *ZTB11*.

### IL-6

Intronic variants rs2069850 and rs34880821 of the *IL-6* gene were found to be significant within the study cohort. These variants have both been referenced before in various publications, but none of which related to endometriosis, or pathways known to be involved. However, the variants have been found to colocalise in the endometriosis cohort, which suggests a potential biological interaction between these variants that are potentially relevant to endometriosis. Data from UCSC (2023) showed rs34880821 to be a Neandertal methylation site which is shown to increase susceptibility to auto-immune/inflammatory conditions (Zhou et al., 2022). Therefore, the reference allele was methylated in Neandertals and Denisovan’s, but this variant abolishes the methylation site, so rs34880821 may exuberate endometriosis development due to the disease being associated with dysregulation of the immune and inflammatory system by aberrant silencing of the area and continuous expression. Specifically, this gene primary function in maturation of B cells and inflammatory pathways (Aliyu et al., 2022). A study conducted by Moghaddam et al., (2022) found dysregulation of *IL-6* to increase angiogenesis and cellular proliferation in the endometrium. This can lead to increased rates of endometrial cell shedding and exacerbate the immune response (Sobstyl et al., 2023). This was also seen in Incognito et al., (2023) study finding increased levels of *IL-6* in endometriosis patients. This probability of heightened *IL-6* expression in endometriosis patients is further validated in research by Benjamin et al., (2020) and Li et al., (2021), finding an increase in *IL-6* expression levels when compared to controls.

The research conducted did not find any significant variants for *TACR3*, but did however, find two variants (rs796412104 and rs565747925) to be consistent in the study population when compared to the GE Population. *TACR3* can be considered as an involved factor in endometriosis development through altered levels of *TACR3* potentially increasing adhesion of endometriotic cells outside of the uterus. This is due to being involved in the P-CSMS which regulates adhesion, vesicle trafficking and adhesion (Verma et al., 2024). This could be further validated by Blasco et al., 2020 finding *TACR3* expression levels to be altered in endometriosis patients’ granulosa cells when compared to controls. Furthermore, *TACR3* is one of the regulators of the HPGA regulating GnRH release (Casteel and Singh, 2020). Due to Xie et al., (2022) finding DEHP exposure to alter expressions of regulators in the HPGA, it is possible that DEHP exposure may increase endometriosis risk through altering TACR3 expression levels.

### The Intersection of Ancient Genetic Variants, Epigenetic Regulation, and Modern Environmental Pollutants in Endometriosis Susceptibility

Our findings highlight how ancient genetic variants inherited from Neanderthals and Denisovans, epigenetic regulation, and modern industrial pollutants may potentially converge to shape population-specific risks for endometriosis. The linkage disequilibrium (LD) analysis of *IL-6* and *CNR1* regulatory variants suggests that evolutionary pressures involving immune regulation and inflammatory responses, when combined with modern environmental exposures, may amplify disease susceptibility (Sankararaman et al., 2014; Harris & Nielsen, 2016).

Neanderthal introgression has significantly influenced *IL-6* regulation, particularly in East Asian populations. The *IL-6* variant rs34880821, located at a Neanderthal-derived methylation site, exhibits strong LD in East Asians (r² = 0.9662, D’ = 1.0), while showing weaker LD in Europeans (r² = 0.5794) and South Asians (r² = 0.8104) (Sankararaman et al., 2014). This suggests that Neanderthal introgression may have played a role in shaping *IL-6* regulatory pathways, particularly in East Asians, where it remains strongly linked. Given that East Asian populations also have the highest reported prevalence of endometriosis (∼15.4%), it is likely that these genetic variants contribute to heightened inflammatory responses, immune dysregulation, and fibrotic lesion formation (Yamamoto et al., 2019; Velarde et al, 2023).

The functionality for the Neanderthal-derived methylation at rs34880821 may indicate another subtle contributor to endometriosis risk. Methylation usually functions as a gene-silencing mechanism, regulating cytokine levels to prevent excessive inflammation (Zeberg & Pääbo, 2020). If methylation is lost, *IL-6* expression could become hyperactive, leading to chronic immune activation, sustaining peritoneal inflammation, fibrosis and deep-infiltrating endometriosis, as *IL-6* is known to drive fibrotic remodelling in reproductive tissues (Velarde et al, 2023). It also may account for heightened neuroinflammation, potentially explaining increased pain sensitivity in East Asian endometriosis patients (Zeberg & Pääbo, 2020).

Similarly, the Denisovan-derived rs806372 variant in *CNR1*, which is involved in immune modulation and pain perception, exhibits moderate LD in East Asians (D’ = 0.8004) but weak LD in Europeans (D’ = 0.4607) and South Asians (D’ = 0.1459) (Sankararaman et al., 2014). Since *CNR1* plays a key role in pain signalling and inflammatory responses, this Denisovan-influenced variant may enhance pain sensitivity in individuals with endometriosis, particularly in East Asian populations (Harris & Nielsen, 2016).

While these ancestral variants may have once provided immune advantages in prehistoric environments, modern environmental factors may be reversing these evolutionary benefits, transforming once-adaptive immune responses into drivers of chronic disease (Velarde et al 2023).

Exposure to endocrine-disrupting chemicals (EDCs), such as bisphenol A (BPA), phthalates, and dioxins, has been shown to demethylate immune regulatory genes, including *IL-6*, leading to excessive cytokine production (Yamamoto et al., 2019). If Neanderthal-derived *IL-6* regulatory variants are already prone to overactivation, additional EDC-induced demethylation could further escalate inflammatory signalling, worsening lesion development. This suggests a gene-environment interaction, where modern industrial chemicals exacerbate genetic predispositions inherited from archaic human ancestors.

In contrast to East Asian and European populations, African populations exhibit significantly lower LD for these *IL-6* and *CNR1* variants (D’ < 0.02, r² < 0.001), suggesting these genetic associations arose post-migration due to Neanderthal and Denisovan introgression (Sankararaman et al., 2014). There is limited available data on the prevalence of endometriosis in African populations, and estimates may vary due to underdiagnosis.

This study supports a novel model for endometriosis susceptibility, in which ancestral genetic variants interact with modern environmental pollutants to modulate immune regulation, chronic inflammation, and pain perception across populations.

## Limitations and future directions

While this study provides novel insights into the genetic and environmental factors influencing endometriosis susceptibility, several limitations must be acknowledged. The sample size was limited, particularly in younger individuals under 29 years old. Additionally, family-based cascade genetic testing was not included, preventing the evaluation of heritability patterns and potential familial aggregation of risk variants. Another limitation is the reliance on medical records to infer endometriosis staging, which introduces the potential for misclassification bias. Furthermore, while this study identified significant associations between regulatory variants and endometriosis risk, functional validation through in vitro and in vivo models is required to confirm their biological relevance.

Future research should focus on expanding the study cohort to include a larger and more diverse population to improve the statistical power of genetic associations and assess whether these findings are consistent across different ethnic backgrounds. Family-based studies incorporating cascade genetic testing could help clarify inheritance patterns and the potential contribution of additional rare variants to endometriosis risk. Further, functional studies are necessary to evaluate how the identified regulatory variants influence gene expression and immune signalling pathways, particularly in response to endocrine-disrupting chemicals. Integrating environmental exposure data with genomic analysis would provide a more comprehensive understanding of how genetic and environmental factors interact in the development and progression of endometriosis. These future directions will be critical for translating genetic discoveries into practical applications for early diagnosis and personalized treatment strategies.

## Conclusion

This study provides a novel perspective on the genetic and environmental interplay driving endometriosis susceptibility, highlighting the influence of ancient regulatory variants and modern industrial pollutants. This research identified statistically significant regulatory variants in *IL-6*, *CNR1*, *IDO1*, and *KISS1R* that may contribute to endometriosis risk through interactions with endocrine-disrupting chemicals. These findings aim to bridge the gap between genetic predisposition, evolutionary selection, and environmental exposures, shedding light on how Neanderthal and Denisovan introgressed variants in immune and pain-regulatory genes may influence disease susceptibility in modern populations.

This is the first study to systematically explore how ancient genetic signatures, epigenetic regulation, and contemporary environmental pollutants intersect in the pathophysiology of endometriosis. These results lay the first steps for future precision medicine approaches, where genetic screening combined with environmental risk assessment may improve early detection and personalized intervention strategies. Moving forward, functional validation of these regulatory variants and their interaction with endocrine disruptors will be crucial in understanding their mechanistic role in endometriosis onset and progression. These findings enhance our understanding of endometriosis as a multifactorial disease but also provide a framework for future research integrating evolutionary genetics, environmental health, and reproductive medicine.

## Data availability statement

Research on the de-identified patient data used in this publication can be carried out in the Genomics England Research Environment subject to a collaborative agreement that adheres to patient led governance. All interested readers will be able to access the data in the same manner that the authors accessed the data. For more information about accessing the data, interested readers may contact or access the relevant information on the Genomics England website: https://www.genomicsengland.co.uk/research.

## Acknowledgements

Posthumous acknowledgement. This work is dedicated to our beloved colleague Dr Elpida Fragkouli for her consistent support, supervision and contribution to the study design and data interpretation.

This research was made possible through access to data in the National Genomic Research Library, which is managed by Genomics England Limited (a wholly owned company of the Department of Health and Social Care). The National Genomic Research Library holds data provided by patients and collected by the NHS as part of their care and data collected as part of their participation in research. The National Genomic Research Library is funded by the National Institute for Health Research and NHS England. The Welcome Trust, Cancer Research UK and the Medical Research Council have also funded research infrastructure.

## Author’s role

A.W. contributed to the study design, collected the data and analysed the data and drafted the manuscript. D.A. contributed to the study design, provided supervision of the study and contributed to the article drafting and editing. D.W. contributed to the study design, analysed the data and editing. J.W. contributed to data gathering. A.M. leads the GE project that this study is a part of, designed the study, data analysis and interpretation, provided supervision of the study and manuscript drafting and editing.

